# Clinical implementation and initial experience with a 1.5 Tesla MR-linac for MR-guided radiotherapy for gynecologic cancer: An R-IDEAL stage 1/2a first in humans/feasibility study of new technology implementation

**DOI:** 10.1101/2021.12.03.21266962

**Authors:** David S. Lakomy, Jinzhong Yang, Sastry Vedam, Jihong Wang, Belinda Lee, Angela Sobremonte, Pamela Castillo, Neil Hughes, Mustefa Mohammadsaid, Anuja Jhingran, Ann H. Klopp, Seungtaek Choi, C. David Fuller, Lilie L. Lin

## Abstract

**Purpose:** Magnetic resonance imaging–guided linear accelerator systems (MR-linacs) can facilitate the daily adaptation of radiotherapy plans. Here, we report our early clinical experience using an MR-linac for adaptive radiotherapy of gynecologic malignancies.

**Methods and Materials:** Treatments were planned with an Elekta Monaco v5.4.01 and delivered by a 1.5 Tesla Elekta Unity MR-linac. The system offers a choice of daily adaptation based on either position (ATP) or shape (ATS) of the tumor and surrounding normal structures. The ATS approach has the option of manually editing the contours of tumors and surrounding normal structures before the plan is adapted. Here we documented the duration of each treatment fraction; set-up variability (assessed by isocenter shifts in each plan) between fractions; and, for quality assurance, calculated the percentage of plans meeting the y-criterion of 3%/3-mm distance to agreement. Deformable accumulated dose calculations were used to compare ATP plans with reference dose plans.

**Results:** Of the 10 patients treated with 90 fractions on the MR-linac, most received boost doses to recurrence in nodes or isolated tumors. Each treatment fraction lasted a median 32 minutes; fractions were shorter with ATP than with ATS (30 min vs 42 min, *P*<0.0001). The y criterion for all fraction plans exceeded >90% (median 99.9%, range 92.4%–100%), i.e., all plans passed quality assurance testing. The average extent of isocenter shift was <0.5 cm in each axis. The accumulated dose to the gross tumor volume was within 10% of the reference plan for all ATP cases. Accumulated doses for lesions in the pelvic periphery were within 1% of the reference plan as opposed to –5.8% to –9.6% for central tumors.

**Conclusions:** The MR-linac is a reliable and clinically feasible tool for treating patients with gynecologic cancer.

## INTRODUCTION

The integration of in-room magnetic resonance (MR) imaging with radiation therapy delivery allows exceptional image-guided treatment that can be readily adapted according to changes in the tumor or surrounding tissues. Because MR imaging provides superior soft tissue characterization over other modalities, treatments can be highly conformal, thereby allowing margin reduction and minimization of inter- and intra-fractional changes in patient anatomy.^1,2^ Despite these apparent advantages, concerns remain regarding resource utilization and overall implementation of MR-guided systems for radiotherapy.

A 1.5 Tesla MR scanner integrated with a linear accelerator was initially proposed as a proof of concept in 2009.^3^ In 2018, the 1.5 Tesla Elekta Unity MR-Linac system (Elekta, Stockholm, Sweden) became the modern commercial incarnation of this design.^2^ Several reports have described implementation of this system for treating tumors at a variety of disease sites, including prostate, gastrointestinal, thoracic, and head and neck.^4-8^ However, little has been reported on its use specifically for gynecologic cancers. Gynecologic cancers may prove particularly suitable for treatment with MR-linacs because the primary and nodal targets are difficult to visualize on CT images, are highly mobile and deformable, are subject to substantial regression throughout treatment, and are susceptible to rotational setup error.^9^

Here we report our initial experiences using a 1.5 Tesla MR-linac to treat gynecologic malignancies, including workflow and feasibility as well as treatment times, quality assurance (QA), and dosimetric variability.

## MATERIALS AND METHODS

### R-IDEAL Approach

For this pilot study, we leveraged the R-IDEAL conceptual framework for technology development in radiotherapy.^10^ Using the R-IDEAL approach, we undertook a structured evaluation of a novel radiotherapy technology/approach, namely, adaptive MR-guided radiotherapy for gynecologic cancer, in a single-institution, prospective implementation series. This assessment included both an “idea” stage (R-IDEAL Stage 1; that is, first-in-human implementation) and a “development” stage (R-IDEAL Stage 2a; that is, reporting technical feasibility “when additional modifications are made to further optimize workflow and technology for innovative treatment delivery.” ^10^ The R-IDEAL conceptual schema allows technology development to be substantiated iteratively as parts of a coherent programmatic project^8,11-15^ and is analogous to the parent IDEAL method for surgery.^16^

The aim of the current study was to demonstrate the first-in-human implementation of adaptive MR-guided radiotherapy (MRgRT) for gynecologic external-beam application (R-IDEAL Stage 1). We further sought to report the technical feasibility of achieving MR-gRT geometric accuracy and the resultant planning and delivery QA within our standard current clinical workflow by using FDA-approved commercial treatment planning systems, reported as a sequential case series (R-IDEAL Stage 2a).

### The MR-linac system

The Elekta Unity MR-linac system (Elekta AB, Stockholm, Sweden) consists of a 7-megavolt (MV) flattening filter free (FFF) beam linac (Elekta AB) integrated with a 1.5 Tesla MR scanner (Philips, Best, The Netherlands).^17^ The linac is mounted on a ring gantry to allow continuous rotation with the 1.5 Tesla magnetic field perpendicular to the entrance of the bore. The source-to-axis distance is 143.5 cm. The maximum field size is 22 cm longitudinally and 57.4 cm laterally. The couch moves only in the longitudinal direction, preventing any non-coplanar beams in treatment planning. The multi-leaf collimator consists of 80 leaf pairs, each with a projected width of approximately 0.72 cm at isocenter.

### Patient selection

This report is a part of the Multi-OutcoMe EvaluatioN of radiation Therapy Using the MR-linac (MOMENTUM) study on behalf of the MR-linac Consortium (NCT04075305),^18^ and was approved by the appropriate institutional review board (IRB PA18-0341). All patients aged 18 years or older who were being treated on the MR-Linac for a primary or recurrent gynecologic malignancy at a single institution were eligible for data extraction. Suitability for treatment on the MR-linac was initially considered by the treating physician and discussed with a multidisciplinary team consisting of physicists, dosimetrists, therapists, and radiation oncologists. Only those patients with gross disease being treated to high doses (>60 Gy EQD_2_) were considered; other considerations included body size (with regard to MR bore width and field-size limitations),^19^ overall mobility (ability to walk to and from the vault), ability to maintain a consistent position over prolonged treatments, and lack of claustrophobia or contraindications to MR imaging (e.g., implanted ferrous metal objects or pacemakers).

### Simulation and treatment planning

Treatments were simulated first on a CT scan followed by an MR scan (ideally obtained on the same day) to facilitate rigid registration (Supplementary Tables E1-E2). Patients were immobilized with a lower-body Vac-Lok system (CIVCO Medical Solutions, Coralville, Iowa, USA) individualized for reproducibility of scan length and treatment site (Supplementary Table E1). The CT scans were obtained with a Philips Brilliance 16-slice CT scanner with ≤2.5 cm slices. The MR scans were obtained with the MR-linac, with 2-minute and 6-minute 3D T1-and T2-weighted sequences acquired via scanning protocols identical to those used during daily MR imaging (Supplementary Table E2). 18F-FDG PET/CT images were obtained as described in published guidelines.^20^

Treatment planning was done with the Monaco treatment planning system, which uses a Monte Carlo–based dose calculation engine (Elekta, Inc., Maryland Heights, MO, USA). Target volumes were contoured on multi-modality images (including T1- and T2-weighted MR and PET/CT images) fused to the CT simulation images. Planning target volume (PTV) margins were typically 3-5 mm around the gross tumor volume (GTV), but were modified as needed for each patient to account for factors such as tumor location and size and receipt of prior radiotherapy. For plans created with the adapt-to-tumor-size (ATS) approach, margins were often reduced to <5 mm (at the physician’s discretion) based on anatomy, proximity to critical structures, and perceived motion. Organs at risk (OARs), that is, bladder, bowel, rectum, pelvic bones, kidneys, and spinal cord, were contoured for each patient. The step-and-shoot intensity-modulated radiation therapy (IMRT) planning technique was used to create MR-linac reference plans. A backup plan for treatment on a conventional linac was also created as a backup for machine downtime.

Pretreatment QA involved verification of monitor units (MU) by using RadCalc (Version 63, Lifeline Software Inc., Austin, TX, USA) and dose measurements by using ArcCheck MR (Sun Nuclear Corp., Melbourne, FL, USA).

### Treatment set-up and delivery

Patients were positioned for treatment delivery according to an index value recorded at simulation to determine their longitudinal position. No external lasers are present within the MR-linac vault.

A 2-minute T2 3D MR imaging scan (Supplementary Table E2) was acquired and subsequently fused via rigid registration with the reference plan image to facilitate verification of daily setup and the need for plan adaptation. Two adaptation workflows were used: adapt to position (ATP) and adapt to shape (ATS).^2^ Briefly, ATP is a workflow wherein an isocenter shift is made on the reference plan based on rigid registration followed by either a dose recalculation or plan reoptimization. The ATS workflow, on the other hand, uses deformable image registration to reproduce contours onto the daily setup image followed by full-plan re-optimization according to daily changes in anatomy. The decision to use ATP vs ATS was left to the physician’s discretion and was generally determined on the basis of extent of tumor shrinkage (by more or less than 3 mm), the need to reduce PTV margins, movement of OARs into the treatment area leading to violation of predefined dosimetric limits, and individual patient tolerance on the day of treatment.

All adaptive plans were then independently checked for MUs by using RadCalc before beam delivery. Real-time motion monitoring was done with orthogonal cine MR imaging during beam-on. After treatment completion, a final quality check was performed, with the stipulation that IMRT QA measurements of the adaptive plan must be within 3% dosimetric difference and 3 mm distance to agreement γ criteria.

### Calculations of accumulated dose

Doses on daily adaptive plans were accumulated for six patients treated with ATP for every fraction. Two types of dose accumulation were compared. For the first, because all ATP plans were done on the reference CT image, we directly summed the fraction dose of each ATP plan to create a summed planned dose, which represents the planned dose for the treatment. In the second type of accumulated dose calculation, daily ATP plan fraction doses were first rigidly shifted to the daily MR coordinate space, and then deformably mapped to the reference CT coordinate space by using an in-house deformable registration tool for dose accumulation.^4^ This accumulated deformed dose accounts for daily variations in anatomy and approximates the actual delivered dose to the target and OARs. The accumulated deformed dose was compared with the summed planned dose by calculating the percentage difference of each plan quality metric with the following equation:

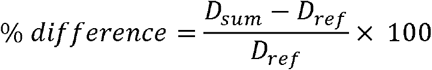

Analyses of targets and nearby OARs were based on the anatomic location of the irradiated lesion and included the GTV and one or more of the following OARs: bladder, femoral heads, bowel, rectum, and sigmoid colon. Doses to the GTV were compared by D_95_, and doses to the OARs were compared by average dose. When a conventional linac system was used to deliver a fraction, only the MR-linac plans were included, and the reference plan was scaled to the number of fractions treated on the MR-linac before calculating the percentage difference.

### Data analysis

Descriptive statistics were used for patient characteristics such as age, body mass index, and performance status (scored according to the Eastern Cooperative Oncology Group [ECOG] criteria).^21^ Extracted treatment characteristics included daily treatment duration, method of adaptation, isocenter shift, and number of beams. Treatment duration was measured from time stamps in Mosaiq, with the total duration measured from the patient entering the vault to beam-off; treatment durations were compared with *t* tests. Daily setup variability was measured from isocenter shift data, which were measured along each X, Y, and Z axis (left-right, superior-inferior, anterior-posterior, respectively) for all adapted fractions.

## RESULTS

### Patients

Ten patients with gynecologic malignancies were treated on the MR-linac system from May 2019 through January 2020 **(Table 1)**. The median age was 67 years (range 40s–80s) and the median body mass index was 26.5 (range 16.5–43.1). All but one patient had an ECOG performance status score of ≤1. Primary gynecologic tumors were cervical (n=4), endometrial (n=3), vaginal (n=1), ovarian (n=1), and peritoneal (n=1); most patients (7 of 10) were being treated for isolated recurrent disease.

**Table 1.** Patient and Treatment Characteristics

| Patient | Age range<br>at<br>Treatment<br>Start | Race | BMI | ECOG<br>PS<br>Score | Primary<br>Tumor<br>Location | FIGO Disease<br>Stage | Tumor Location | Chemotherapy | MRL<br>Use | Total<br>MRL<br>dose,<br>Gy | No. of<br>Fractions | Adaptation<br>Method | No. of<br>Beams |
| --- | --- | --- | --- | --- | --- | --- | --- | --- | --- | --- | --- | --- | --- |
| 1 | 41-45 | Black | 20.6 | 0 | Cervix | R | Vaginal cuff | Cisplatin | Boost | 16 | 8 | ATP | 9 |
| 2 | 76-80 | White | 32.8 | 1 | Uterus | R | Inferior splenic node | Carboplatin+<br>paclitaxel | Full<br>plan | 61.6 | 28 | ATP/ATS | 7 |
| 3 | 66-70 | Unknown | 43.1 | 1 | Uterus | R | Left external iliac node | Carboplatin+<br>paclitaxel | Full<br>plan | 40 | 5 | ATP | 6 |
| 4 | 56-60 | White | 16.5 | 1 | Cervix | IIIB | Cervical | Cisplatin | Boost | 22 | 8 | ATP/ATS | 8 |
| 5 | 46-50 | Asian | 22.3 | 0 | Cervix | R | Left pelvic sidewall | None | Full<br>plan | 42.5 | 5 | ATP | 7 |
| 6 | 66-70 | White | 31.9 | 1 | Ovary | R | LEI and left pelvic mass | None | Boost | 16 | 8 | ATP/ATS | 7 |
| 7 | 46-50 | White | 21.3 | 0 | Cervix | R | Left Ischium | None | Full<br>plan | 27 | 3 | ATP | 8 |
| 8 | 66-70 | White | 18.9 | 2 | Peritoneum | R | Left pelvic/abdomen | Cisplatin | Boost | 30 | 15 | ATP/ATS | 9 |
| 9 | 66-70 | Hispanic | 30.7 | 1 | Vagina | I | Vaginal cuff | Cisplatin | Boost | 10 | 5 | ATP | 9 |
| 10 | 86-90 | Black | 30.6 | 1 | Uterus | IIIC | Uterine fundus | None | Boost | 12 | 5 | ATP | 11 |

Abbreviations: BMI, body mass index; ECOG, Eastern Cooperative Oncology Group; PS, performance status; R, recurrent; ATP, adapt-to-tumor-position; ATS, adapt-to-tumor-shape; LEI, left external iliac node.

The clinical presentations in this analysis varied. Patients 1, 4, 6, 8, 9, and 10 received central pelvic irradiation on the MR-linac to either the whole pelvis or to centrally located lesions such as the uterine fundus; patients 3 and 5 were treated to peripheral pelvic lesions such as the external iliac node and pelvic sidewall lesion, and the others were treated to extrapelvic areas such as an infrasplenic node (patient 2) and the left ischium (patient 7). In 6 patients, the MR-linac was used to deliver boost doses in a hybrid conventional linac–MR-linac treatment regimen; the other 4 patients were treated exclusively with the MR-linac.

### Treatment characteristics

A total of 92 fractions were planned, 90 of which were to be delivered on the MR-linac; the 2 missed fractions were delivered with a conventional linac owing to unexpected MR-linac system downtime. Six patients received concurrent platinum-based chemotherapy (40 mg/m^2^ cisplatin for four, carboplatin-paclitaxel for two).

The median total dose delivered on the MR-linac was 22.5 Gy (range 10.2–60.5 Gy), in a median of 6 fractions (range 3–28); the median dose per fraction was 2.2 Gy (range 2–9 Gy). The median number of beams per plan was 8 (range 6–11). All fractions passed IMRT QA with a y-value in excess of 90% (median 99.9%; range 92.4%–100%), indicating a high level of agreement between expected and delivered dose distributions (Supplementary Figure E1).

The average isocenter shift for daily patient setup was <0.5 cm in the X, Y, and Z axes (left-right, superior-inferior, and anterior-posterior, respectively). The absolute maximum isocenter shift in any fraction was 3.5 cm for the X axis, 3.3 cm for the Y axis, and 1.9 cm cm for the Z axis (Supplementary Figure E2). A large (>3 cm) shift was considered necessary for one patient because of the laterality of disease and limits on the MR field of view.

Of the total 90 fractions, 73 were delivered with the ATP method and 17 were delivered with the ATS method; no patient was treated solely with the ATS method. The median duration of treatment was 32 minutes overall and was significantly shorter with ATP (median 30 min, range 17–66 min) than with ATS (median 42 min, range 30–65 min), P<0.0001 (Supplementary Figure E3).

### Dose accumulation analysis

The accumulated deformed dose for the GTV was within 10% of the summed planned dose for all patients treated with the ATP method **(Figure 1)**. For patients whose irradiated target lesions were within the periphery of the pelvis or a fixed bony lesion, the relative GTV dose was within 1%; by contrast, the relative GTV dose for centrally located lesions ranged from –5.8% (uterine fundus boost) to –9.6% (vaginal cuff boost) of the summed planned dose. The relative dose calculations for OARs varied, with that to the femoral heads all being within 3% and that to the bladder and rectum being within 20%. Relative disagreement between accumulated deformed doses and summed planned doses was greatest for the bowel, rectum, and sigmoid colon, with some cases exceeding 20%.

**Figure 1.**
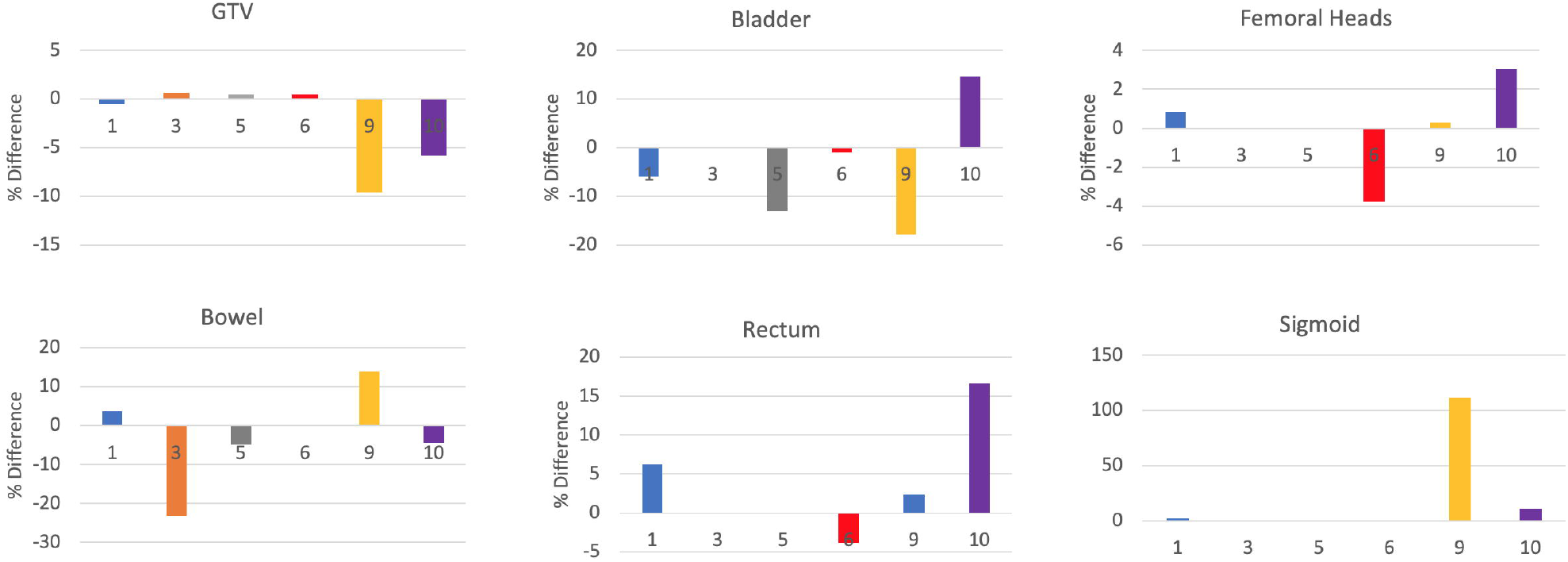
% differences between accumulated deformed dose and summed adaptive plan dose to GTV & OARs]. Differences between the accumulated deformed dose and the summed adaptive plan dose, shown for the gross tumor volume (GTV) and various organs at risk (OARs) for 6 patients treated with solely ATP plans. Positive values indicate that the accumulated deformed dose exceeded the summed plan dose. Doses to the GTV are expressed as D_95_ (the dose to 95% of the volume); doses to organs at risk are expressed as mean dose. The numbers on each bar refer to the patients as shown in Table 1. The various GTV targets for each patient were as follows: Patient 1, the recto-vaginal space; Patient 3, external iliac node; Patient 5, pelvic sidewall; Patient 6, left ischium; Patient 9, vaginal cuff boost; and Patient 10, uterine fundus. If an OAR was not considered in a particular patient, then no bar appears on the panel (e.g., for Patient 3, the bladder, femoral heads, rectum, and sigmoid colon were not considered as OARs).

## DISCUSSION

Advances in radiation therapy for gynecologic malignancies have led to increasingly conformal treatments and image-guided adaptive targeting over time, from the widespread adoption of CT-planned external-beam methods^22,23^ to the development of MR imaging–based image-guided adaptive brachytherapy.^24,25^ MR-guided adaptive [external-beam therapy represents the next step in this advancement. Rather than relying on a single CT data set that may not reflect the geometry of the target and OARs at the time of treatment delivery, the MR-linac system allows daily adaptation to plans that account for changes in the highly mobile deformable structures within the pelvis **(Figure 2)**. This report is among the first to provide details on the feasibility and initial clinical experience with using a 1.5 Tesla MR-linac specifically for patients with gynecologic cancer.

**Figure 2.**
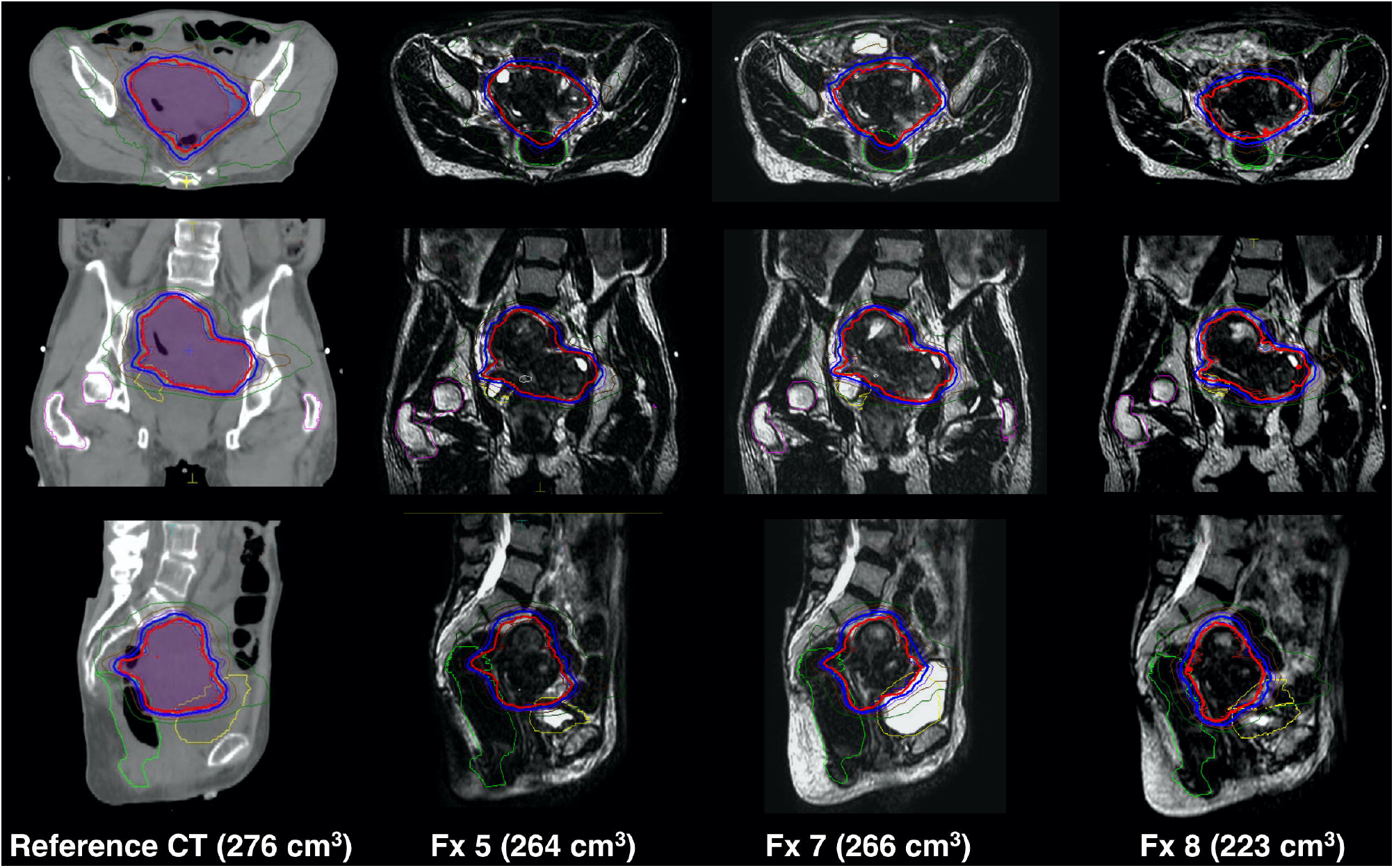
CT reference plan and adapt-to-shape plans throughout treatment. Comparisons of CT-based reference scans (left column) with magnetic resonance (MR) images of daily adapt-to-tumor-shape plans at fractions 5, 7, and 8 for Patient 6, a woman in her 60s with recurrent ovarian cancer treated to 50 Gy with a conventional linear accelerator (linac) (not shown) and given a sequential 16-Gy pelvic boost with the MR-linac. Top row, axial views; middle row, coronal views; and bottom row, transverse views. The gross tumor volume (GTV) is outlined in in red; the planning target volume (PTV; i.e., GTV + a 2-mm expansion) in blue; the bladder in yellow; and the rectum in green. The GTV on the reference CT scan was 275 cm ^3^; volumes on the daily adaptation scans were 264 cm^3^ at fraction 5, 266 cm^3^ at fraction 7, and 223 cm^3^ at fraction 8. These scans illustrate the high level of conformality possible with the ability of the MR-linac system to account for differences in bowel and rectal filling between daily fractions.

The best basis for identifying patients who would derive the greatest benefit from MR-linac treatment remains unclear. In this study, patients were selected for MR-linac treatment if they had gross nodal or primary disease that was either mobile or closely approximating OARs such as the bladder, rectum, or bowel; in such cases, daily imaging would help to reduce treatment margins and reduce the dose to OARs. The Elekta Unity system used for this study has a structural limit of a 22-cm longitudinal field size, thus precluding treatment for patients with extensive nodal or metastatic disease requiring an extended field. However, methods for treating larger fields are being actively investigated. Most of the patients in our study were given isolated nodal or tumor boost doses with the MR-linac system.

One significant concern regarding the use of MR-linac systems is the time required to deliver each treatment fraction; in this study, the median fraction time was 32 minutes, with a range of 17 to 66 minutes; notably, the ATS plans took longer, at a median 42 minutes. Although these values are similar to the fraction times for treating head and neck cancer (median 41 minutes) or pelvic nodal disease and prostate (30-40 minutes) with the Elekta Unity,^4,5,26^ they are much longer than the typical 3-4 minutes of “beam-on” time within 15-minute scheduled blocks for patients receiving conventional IMRT. That said, all adaptive plans in the current study passed QA checks, with y exceeding 90% (median 99.9%, range 92.4%–100%), indicating a high level of agreement between expected and delivered dose distributions. The high pass rate indicates good agreement between expected and delivered dose distributions; indeed, this dosimetric stability is probably attributable to the comprehensive series of safety checks that are implemented at each level.

The average isocenter shift was <0.5 cm in each axis. Although the magnitude of isocenter shifts was larger in this study than in another of patients with head and neck cancer,^8^ this could be explained by the use of a stabilizing mask used for head and neck cases and the greater degree of inter- and intra-fractional movement of pelvic and abdominal lesions in the current study. Notably, these isocenter shifts should be interpreted as couch shift corrections and should not be used to derive margins, for the following reasons. For one thing, the set-up accuracy of the MR-linac for these patients is typically not as accurate as regular IGRT because external lasers are not used in MR systems. For another, the daily adaptation possible with the MR-linac accounts for the isocenter shift during the online adaptive planning. Further studies of marginal status would consider the residual match due to anatomic change or deformation set-up error, with individual evaluations of different target locations. Given the small number of patients and the heterogeneity in target locations in the current study, we did not have enough data to draw any conclusions regarding appropriate margins.

Our dose accumulation analysis showed that for ATP plans (that is, plans adapted according to tumor position as opposed to tumor shape), the differences between summed planned dose and accumulated deformed dose varied significantly for different targets. Peripheral lesions and bony lesions (i.e., fixed lesions with little mobility) had only minor differences between planned and deformed dose relative to central pelvic lesions, for which the dose difference was often >5%. For example, for patient 9, who was treated with a vaginal cuff boost, the difference between planned and deformed dose was – 9.8%; no doubt the location of the lesion (adjacent to the bladder) meant that daily variability in bladder filling resulted in registration challenges and thus a higher degree of plan disagreement. Our observations should be confirmed in larger studies. Another study in which the same in-house tools as described here were used for prostate cancer found differences of <5% in prostate volumes,^4^ which probably reflects the relatively immobility of prostate volumes relative to pelvic and abdominal targets. We also found significant differences in agreement among the various OARs, which may represent the limitations of ATP plans in simply carrying over the original contours of the initial simulation scan. Although the ATS approach probably results in an actual delivered dose that is more akin to the planned dose, this method does increase the duration of treatment.

In summary, we found that the Elekta Unity 1.5 Tesla MR-linac is a versatile system that can be used to treat gynecologic disease at a variety of sites, alone or in combination with other radiation techniques in a reliable, effective manner. Treatment duration is acceptable when compared with that of conventional radiation modalities, especially when ATP is used. Although our study was limited by the small cohort and the heterogeneity of treatments, it does show the initial feasibility of using this novel modality in gynecologic oncology. Further exploration is required to assess how the MR-linac might be implemented for patients requiring extended-field treatments; for patients for whom brachytherapy is unavailable, and how the MR-linac might replicate similar doses; and for how current margin standards could be safely attenuated given the enhanced adaptability of the system. Thus, even though the MR-linac is a novel treatment modality, we found that MR-linac treatment plans were clinically similar to the current standard of care, were well-tolerated, and allow greater precision and individualization in the treatment of gynecologic malignancies.

## Data Availability

Research data are stored in an institutional repository and will be shared upon reasonable request to the corresponding author.

## Acknowledgments

The authors thank Christine F. Wogan, MS, ELS, from MD Anderson’s Division of Radiation Oncology for editorial assistance.

