## Supplemental for "Clinical implementation and initial experience with a 1.5 Tesla MR-linac for MR-guided radiotherapy for gynecologic cancer: An R-IDEAL stage 1/2a first in humans/feasibility study of new technology implementation"

**SUPPLEMENTARY MATERIAL**

**Supplementary Table E1**. Requirements for Treatment Simulations

| Body Positioning | Supine |
| --- | --- |
| Head positioning | Neutral |
| Arm positioning | On chest, above chest if treating upper abdomen |
| Other positioning |  |
| Patient positioning devices | Lower Vac-Lok; Upper Vac-Lock if treating upper abdomen. Index bars along couch to reproduce immobilization aid location. |
| Bolus | None |
| CT (PET/CT, other) extent | Individualized for treatment area. For pelvic sites, generally mid-lumbar spine to mid-femur. CT in the same position is required for dose calculation, and should use comparable slice thickness reconstruction to the MR-linac-simulation scan. |
| CT (PET/CT, other) slice thickness, mm | ≤2.5 |
| Contrast | None |
| 4D extent and goals of assessment | None |
| Isocenter location | Within the target volume |
| Other | For ATL2/Unity current version, the maximum superior-inferior treatment field length is 22 cm, thus necessitating a maximum acceptable superior to inferior distance of ≤21 cm (≤20 cm preferred) after planning target volume expansion.  A tabletop overlay is used on the CT scanner to mirror MR-Linac couch top. A hood-shaped dome is also used to represent the MR-linac’s maximum clearance and to ensure fit onto the MR-Linac.  Ideally, MR- and CT-based treatment simulation should occur within <24 hours; intervals between CT- and MR-based simulation scans longer than 7 days should be discouraged.  Contrast (CT or MRI) was not routinely used |

**Supplementary Table E2.** MR-Linac Sequences

|  | **T2w,**  **2 min** | **T1w,**  **2 min** | **T2w,**  **6 min** | **T1w, 6 min** |
| --- | --- | --- | --- | --- |
| MR manufacturer | Philips | Philips | Philips | Philips |
| Field strength | 1.5 T | 1.5 T | 1.5 T | 1.5 T |
| Sequence name | T2 3D Tra 2min | T1 3D Tra 2min | T2 3D Tra | T1 3D Tra |
| Coil | Multi coil | Multi coil | Multi coil | Multi coil |
| Slice thickness, mm | 2 | 2 | 2.2 | 2.4 |
| Resolution X/Y plane (reconstructed), mm | 0.83 | 0.83 | 0.68 | 0.68 |
| Slice gap, mm | 1 | 1 | 1.1 | 1.2 |
| Bandwidth, Hz/pixel | 740 | 432 | 459 | 344 |
| FOV, mm | 400 | 400 | 520 | 380 |
| No. of slices | 300 | 300 | 264 | 208 |
| Time to echo, ms | 277.818 | 4.604 | 375 | 4.501 |
| Time to repetition, ms | 1535 | 11 | 2100 | 13 |
| Flip angle, degrees | 90 | 30 | 90 | 27 |
| Slice acquisition | 3D | 3D | 3D | 3D |
| Sequence type | SE | GR | SE | GR |

Abbreviations: MR, magnetic resonance; FOV, field of view; SE, spin echo; GRE, gradient echo

**Supplementary Figure E1.** Intensity-Modulated Radiation Therapy (IMRT) Quality Assurance (QA) Measurements for Adaptive Plans

| Pass Rate % | No. of MRL Plans |
| --- | --- |
| 92 | 1 |
| 93 | 0 |
| 94 | 0 |
| 95 | 0 |
| 96 | 1 |
| 97 | 7 |
| 98 | 16 |
| 99 | 21 |
| 100 | 44 |

The $\gamma$ criteria of 3% dose/3-mm distance to agreement was measured with ArcCheck. MRL, magnetic resonance imager-linear accelerator system.

**Supplementary Figure E2**. Magnitude and Direction of Isocenter Shift Changes

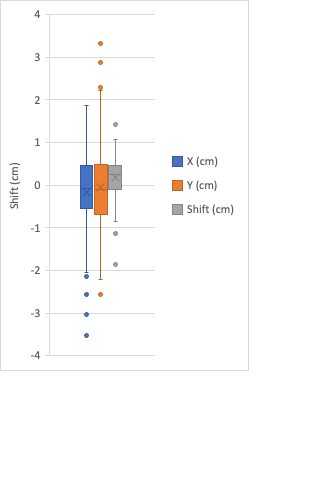

X refers to left/right axes, Y to superior/inferior axes, and Z to anterior/posterior axes

**Supplementary Figure E3.** Treatment fraction duration based on the adaptation-to-tumor-position (ATP) method and the adapt-to-tumor-shape (ATS) method.

ATP

Percent of Fractions

| Duration (minutes) | ATP |
| --- | --- |
| 0-9 | 0 |
| 10-19 | 7.8125 |
| 20-29 | 42.1875 |
| 30-39 | 42.1875 |
| 40-49 | 3.125 |
| 50-59 | 3.125 |
| 60-69 | 1.5625 |

ATS

Percent of Fractions

| Duration (minutes) | ATP |
| --- | --- |
| 0-9 | 0 |
| 10-19 | 0 |
| 20-29 | 0 |
| 30-39 | 43.75 |
| 40-49 | 31.25 |
| 50-59 | 18.75 |
| 60-69 | 6.25 |
